# Social-spatial network structures among young urban and suburban persons who inject drugs in a large metropolitan area

**DOI:** 10.1101/2023.02.21.23286255

**Authors:** Qinyun Lin, Javier Andres Rojas Aguilera, Leslie D. Williams, Mary Ellen Mackesy-Amiti, Carl Latkin, Juliet Pineros, Marynia Kolak, Basmattee Boodram

## Abstract

**Background:** It is estimated that there are 1.5% US adult population who inject drugs in 2018, with young adults aged 18-39 showing the highest prevalence. PWID are at a high risk of many blood-borne infections. Recent studies have highlight the importance of employing the syndemic approach to study opioid misuse, overdose, HCV and HIV, along with the social and environmental contexts where these interrelated epidemics occur in already marginalized communities. Social interactions and spatial contexts are important structural factors that are understudied.

**Methods:** Egocentric injection network and geographic activity spaces for young (aged 18-30) PWID and their injection, sexual, and social support network members (i.e., where reside, inject drugs, purchase drugs, and meet sex partners) were examined using baseline data from an ongoing longitudinal study (n=258). Participants were stratified based on the location of all place(s) of residence in the past year i.e., urban, suburban, and transient (both urban and suburban) to i) elucidate geospatial concentration of risk activities within multi-dimensional risk environments based on kernel density estimates; and ii) examine spatialized social networks for each residential group.

**Results:** Participants were mostly non-Hispanic white (59%); 42% were urban residents, 28% suburban, and 30% transient. We identified a spatial area with concentrated risky activities for each residence group on the West side of Chicago where a large outdoor drug market area is located. The urban group (80%) reported a smaller concentrated area (14 census tracts) compared to the transient (93%) and suburban (91%) with 30 and 51 tracts, respectively. Compared to other areas in Chicago, the identified area had significantly higher neighborhood disadvantages (e.g., higher poverty rate, *p*<0.001). Significant (*p*<0.01 for all) differences were observed in social network structures: suburban had the most homogenous network in terms of age and residence, transient participants had the largest network (degree) and more non-redundant connections.

**Conclusion:** We identified concentrated risk activity spaces among PWID from urban, suburban, and transient groups in a large outdoor urban drug market area, which highlights the need for considering the role of risk spaces and social networks in addressing the syndemics in PWID populations.

## 1. Introduction

It is estimated that there were around 3.7 million people who inject drugs (PWID) in the U.S. in 2018 (Bradley et al., 2022), representing a 5-fold increase since the previous estimate from 2011 (Lansky et al., 2014). The 3.7 million PWID accounts for 1.5% of all adult population, with young adults aged 18-39 showing the highest prevalence (Bradley et al., 2022). PWID are at a high risk of many blood-borne infections (CDC, 2021). Indeed, IDU is the most reported primary risk factor among hepatitis C (HCV) cases in the United States (67%, according to CDC). Such emerging patterns, however, are embedded in the syndemic of opioid misuse, overdose, HCV, and HIV, along with the social and environmental contexts where these interrelated epidemics occur in already marginalized communities (e.g., Friedman et al., 2016; McLean, 2016; Mizuno et al., 2015; Singer & Clair, 2003; Storr et al., 2004). Using the syndemic approach, we emphasize that each health condition (e.g., overdose, HIV, HCV) interacts with each other “at the levels of causes, consequences, and needed responses” (Perlman & Jordan, 2018). In such a sense, individual engagement in risk practices such as syringe sharing impacts all downstream adverse health outcomes. More importantly, unlike the co-morbidity concept, the syndemic approach focuses on the “coinfection and synergistic interaction of diseases of social conditions,” (Singer & Clair, 2008) highlighting the importance of examining structural level factors in communities and neighborhoods. Using the socio-ecological framework in the social determinants of health (SDOH) literature, such structural-level factors include social interactions at the interpersonal level (e.g., injection risk networks), as well as equitable access to social, economic, health care, physical, or built environmental conditions at the neighborhood level (e.g., access to harm reduction, unstable housing, community policy, drug quality).

Many prior studies have examined the role of such structural-level factors on IDU risk behaviors that impact health outcomes and downstream infections. For example, social network characteristics like social norms are shown to be associated with risk behaviors such as the sharing of syringes and drug preparation equipment (De et al., 2007; Latkin et al., 2010). Highly cohesive and centralized networks are also demonstrated to be facilitative of HIV transmission (Yong et al., 2013). On the other hand, some other research examines the association between health and risk behaviors and place-specific features at the neighborhood level, such as socioeconomic vulnerability, primary and specialist clinical providers and alcohol outlet density (e.g., Chen et al., 2019; Kolak et al., 2020). There is also evidence that perceived neighborhood disadvantage strongly relates to risk behaviors and IDU (Latkin et al., 2007, 2009).

As implied by the socio-ecological model, social interactions are embedded in specific geographic locations, meaning the social network effects at the interpersonal level interact with the other SDOH conditions at the community level to impact risk behaviors and health outcomes. On one hand, social network members may spatially bridge high and low HCV prevalence locations (e.g., urban vs. suburban or different public spaces where people inject drugs) through mobility patterns and transience (Boodram et al., 2018; Wylie et al., 2007). On the other hand, geographic locations may influence social network characteristics and structure. For example, public injection spaces may promote the formation of high-risk IDU networks (Tempalski & McQuie, 2009). Altogether, socio-structural influences not only impact individual behaviors but can also concentrate adverse health outcomes within individuals’ networks as well as embed these social networks within places with limited resources (Brawner et al., 2022). For instance, within defined spaces, studies have found spatial clustering of risk behaviors and norms among drug use and sexual partners (Latkin et al., 2007) that may be related to perceived “neighborhood disorder,” defined as the clustering of violence, housing problems, economic stress, and drug market activities (Latkin et al., 2013). More importantly, recent work highlights the legacy role of laws and practices driving and reinforcing racial health inequalities through processes like residential segregation, housing policies, and other aspects of institutional racism (see review by Brawner et al., 2022). Moreover, variation in neighborhood characteristics could theoretically influence the behaviors of PWID through multifaceted, interconnected but systematic mechanisms (Tempalski & McQuie, 2009). Examples include substance use policies and their enforcement by police, availability of related services like syringe service programs (SSPs) and drug treatment programs, population density, income and poverty, the characteristics of drug supply chains and markets (e.g., drug quality), access to public transportation, as well as proximity to main thoroughfares (Rhodes et al., 2005, 2006).

Given the theoretical importance of place in understanding substance use behaviors, we employ the concept of activity space to both examine locations of risk behaviors and understand how PWID interact with their environment (Martinez et al., 2014). Defined as the geographic extent in which people undertake their routine activities (Ren, 2016), activity spaces include all relevant locations including the place of residence and risk behaviors for PWID. As a special case of how the place may interact with social networks to impact risk behaviors, the place of residence may shape PWID’s social networks as well as their locations of risk behaviors. As such, we consider the potential heterogeneity in terms of both social network characteristics and their locations of risk behaviors among different PWID groups based on their recent places of residence. To highlight the structural factors at the community level, we then describe the neighborhood characteristics of those geographically overlapped spaces where different PWID groups’ risk behaviors concentrate, which we also call as the concentrated risk activity space (CRAS). In Figure 1 we summarize such conceptual framework for this study. To the best of our knowledge, this is the first study that simultaneously examines geographic variation in residence, characteristics of social networks, and neighborhood conditions where risk behaviors occur among PWID. Understanding these interconnected factors could provide valuable insight for the development of multilevel interventions (e.g., distribution of SSPs, HIV/HCV testing sites, drug treatment centers) to address the syndemic of opioid misuse, overdose, and other downstream infectious disease transmissions such as HCV and HIV.

**Figure 1.**
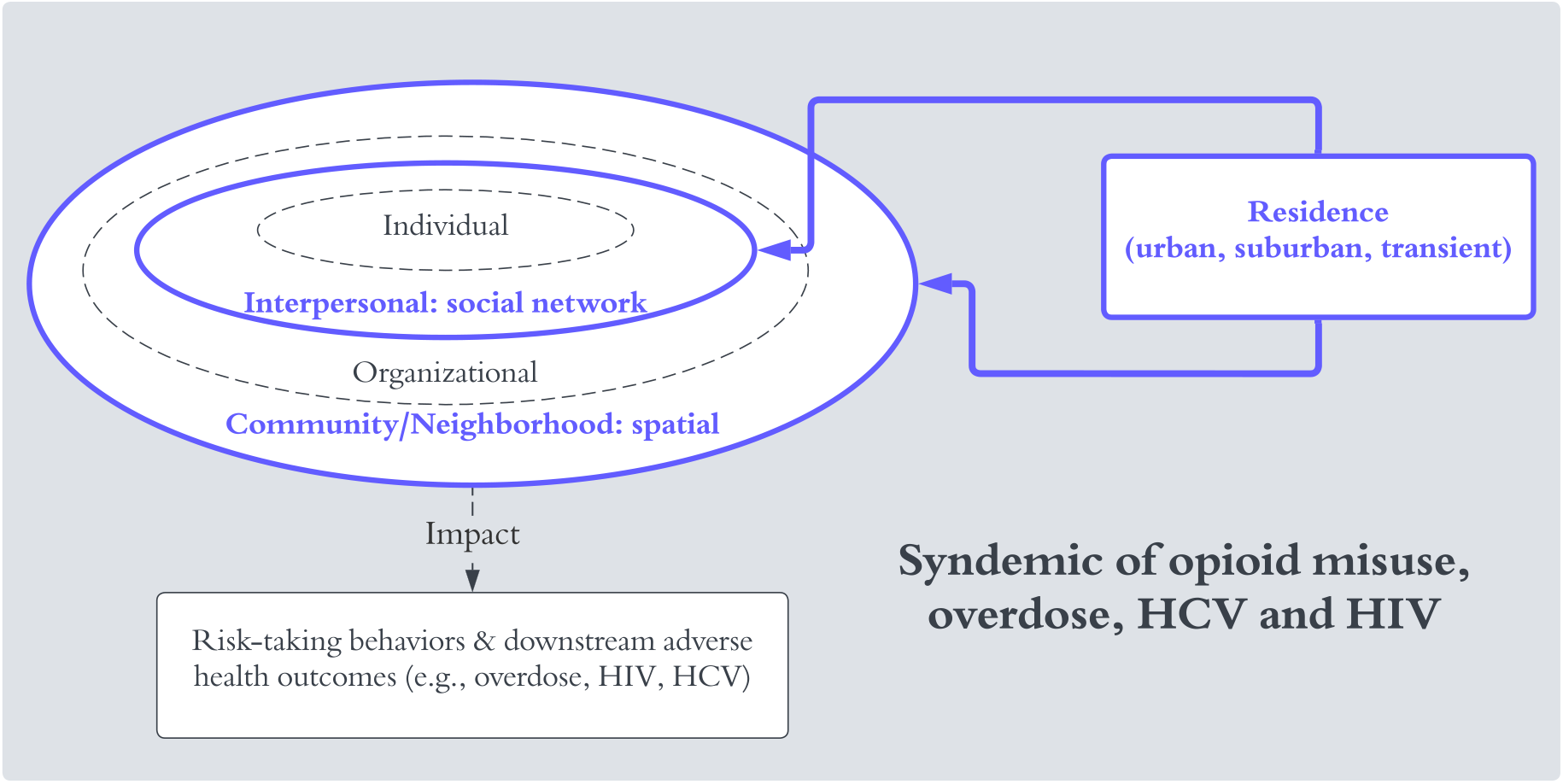
Conceptual framework of the study.

## 2. Methods

As we discussed before, our goal is to examine how different PWID residential groups have similar or different experience in the syndemic of opioid misuse, overdose, and downstream health outcomes so that we can have a more comprehensive understanding of the multidimensional risk environment as described by Rhodes (2002). To achieve this goal, we adopt an exploratory and descriptive approach. We first identify PWID residential groups based on their most recent residence. After that, we characterize each group by examining their individual attributes such as demographic and travel behaviors, social network characteristics, and the space where risk activities occur. Kernel density estimates (KDE) are used to identify those risk activity spaces (CRASs). The identified CRASs are then spatially joined to census tracts so that these areas could be further contextualized by neighborhood characteristics, using other census tracts in Chicago as a reference. To provide a direct, straightforward description of the multidimensional risk environment across different levels for PWID experiencing the syndemic, we intentionally use bivariate analyses instead of multivariate regression analyses in this study. In Figure 2, we summarize the analytical process.

**Figure 2.**
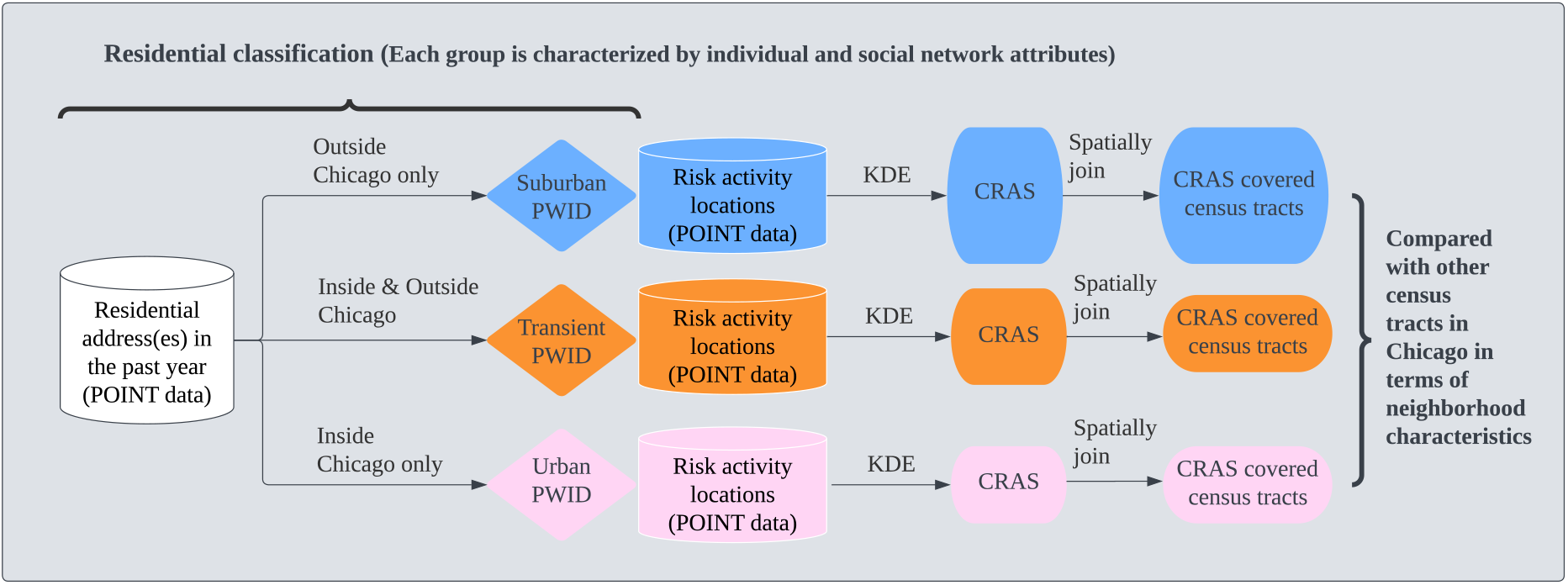
Overview of analytical process.

### 2.1 Recruitment, data collection, and sample

Our study used baseline data from a longitudinal network-based study of young PWID (aged 18– 30) and their network members. Recruitment, enrollment, and data collection methods have been previously described (Mackesy-Amiti et al., 2022). Importantly, multiple recruitment approaches have been utilized to ensure a diverse sample of urban and suburban PWID. In particular, participants were recruited from the SSPs at two field sites of a community outreach center in Chicago, in outdoor drug market areas using an outreach van, fliers posted at community-based organizations serving PWID, and through social media online advertisements. The SSPs and the outdoor drug market areas, while located in urban areas, attract urban and suburban PWID giving easy access via transportation (public and driving). To be eligible for the study, all ego participants had to be current injectors (i.e., at least inject once in the past 30 days) and had to be willing to help recruit their injection network members (alters) for second wave of data collection. See Figure 1 in Mackesy-Amiti et al 2022 for more details of the sampling process. This study was reviewed and approved by the IRB Board at University of Illinois at Chicago (Protocol #2017-0388).

All participants completed a baseline computer-assisted interviewer-administered questionnaire on demographic background, substance use, injection and other risk behaviors, HIV and HCV testing history, and their drug injection experience. Participants also completed (i) an egocentric (versus sociocentric) network inventory and survey on their social networks in the past 6 months using GENSI software (Stark & Krosnick, 2017) and (ii) a geographic survey on their risk activity spaces in the past year using software linked to Google Maps that were geocoded using the OpenCage Geocoding API. All participants received rapid HIV and HCV antibody testing (Orasure Technologies, 2010) was conducted to determine prevalence of exposure (past or current infection for HCV). In addition to collecting data directly from participants, we also collected data from publicly available data sources on structural characteristics of neighborhoods that may relate to PWID’s risk behaviors (see Measures section below).

During October 2018-March 2021 we recruited 295 PWID. Since this study focuses on urban and suburban PWID in Chicago area, we excluded participants who did not provide any residential addresses or lived in the past year outside the state of Illinois, resulting in a final analytical sample of 258 individuals with 590 reported residential addresses and 2572 reported risk activities in the past year.

### 2.2 Measures

#### 2.2.1. Sociodemographic characteristics

Participants self-reported gender (male, female, or transgender), age, race, Hispanic/Latinx ethnicity, and days homeless in the past 6 months. Race and ethnicity were combined to create an indicator variable with categories including non-Hispanic white, non-Hispanic Black, Hispanic, and non-Hispanic other race/ethnicity. Non-Hispanic Black PWID were grouped with other non-white non-Hispanic individuals in the “Others” group due to the small number.

#### 2.2.2. Injection risk behaviors

Participants were asked to indicate drugs used (injection and non-injection) in the past six months. Injection risk behavior frequency was assessed on a 5-point Likert-type scale (never, less than half the time, about half the time, more than half the time, or always). Questions assessed receptive syringe sharing (RSS) (“When you shot-up in the last 6 months, how often did you use a syringe that you know for sure had been used before by someone else?”), equipment sharing (“When you injected drugs in the last six months, how often did you use any of the following items with other people? (a) drawn from the same cooker, (b) used the same cotton, (c) used the same rinse water”), and backloading (“In the last six months, how often did you inject with a syringe AFTER someone else has squirted drugs into it from their syringe?”). Each of these measures was dichotomized to indicate any of the specified behavior in the past six months. Participants also reported the approximate date of their last overdose, which was used to compute any overdose in the past six months.

#### 2.2.3. Social networks

We follow egocentric network design (versus sociometric). For each ego, we asked them about their network members, including characteristics of each alter and the social connections between different alters. Although such design may not provide information regarding how different egos’ networks are connected, it has gained much popularity in public health given “its focus on individuals, groups and communities (Djomba & Zaletel-Kragelj, 2016).” The egocentric approach is also appropriate for studying bloodborne infectious disease transmission risk, the focus of this study, given that one’s immediate network connections are more relevant to the individual’s risk for infection. Moreover, to capture PWID’s network exposure from different experiences, we considered three types of networks in the past six months, namely injection network, sexual network, and social support network. The injection network refers to alters with whom the ego has injected drugs. The sexual network includes alters with whom the ego has sex. The social support network refers to alters who provided the ego with personal support by providing advice and/or let the ego stay at their place if needed. For each ego, different networks may overlap with each other. This means the same alter could be listed in more than one network of the ego when, for example, the alter and ego both injected drugs together and provided support. Summary measures were computed for each network using NetworkX version 2.4 (Hagberg et al., 2008) as described in detail in a previous publication (Mackesy-Amiti et al., 2022). These measures quantify each ego’s network structure through network size, closeness and characteristics of network members, which could also be viewed as additional attributes or characteristics for each PWID. In Table 1 we present definitions for each measure used in this paper.

**Table 1.** Definitions of social network measures used in the study

| Measure | Definition |
| --- | --- |
| Network Degree | The number of network members an ego has. |
| Mean strength of ties | The average strength of ego-alter tie. |
| Age standard deviation of alters | The standard deviation of all alters' ages. |
| Average age of alters | The average age of all alters. |
| Max age of alters | The age of the oldest alter. |
| % alters living in Cook county | The percentage of alters living within Cook County |
| Residence homophily | The percentage of an ego's alters that live in the same area as the ego. |
| Network effective size | A measure of non-redundant connections. If the ego is connected to alters which are in turn connected with each other, there is a greater redundancy, and thus the effective size will be smaller. A high effective size is thus indicating high richness in structural holes (Burt, 1992; Latora et al., 2013). |
| Network tie density | The number of ties in the network divided by the number of possible ties. |

#### 2.2.4. Activity spaces

The activity spaces of each participant were converted to spatial point data using geographic coordinates from the corresponding geographic survey. While activity space research often focuses on movement data to represent activity spaces across higher temporal resolution, we were limited to focusing on specific locations of interest as they were self-reported by participants. While we did not have explicit movement data, survey questions did provide insight into transportation behaviors. Activity spaces thus included places where participants would purchase drugs, inject drugs, hang out, or meet sex partners. In this analysis, we focus on the interaction of injection and sexual activities in risk activity spaces.

#### 2.2.5. Residential classification

Our prior work showed significant differences in risk based on region of residence of PWID in the past year (Boodram et al., 2010, 2015, 2018). Therefore, we classified participants similarly in these analyses. Of note, many individuals reported more than one residential address in the past year. If all reported residential locations were inside Chicago, the individual was classified as urban; on the contrary, if all locations were outside Chicago, they were labeled as suburban. Participants who reported residential locations both inside and outside the Chicago city were labeled as “crossover” transient participants. Such classification serves as a tool to understand the overlapping and divergent patterns among individuals of each stratum in terms of their characteristics of risk activity space (i.e., places where they purchase, inject drugs, and met sex partners) and social networks.

#### 2.2.6. Neighborhood charactseristics

Measures of both physical and social disorder of neighborhoods (Latkin et al., 2009; Marco et al., 2015) were obtained from publicly available data sources at the census tract scale to characterize concentrated risk activity spaces. Physical disorder measures obtained include percentage of vacant housing (ACS, 2018), percentage of long-term occupancy (ACS, 2018), foreclosure rate (HUD, 2009) and traffic volume (IDOT, 2019). Social disorder measures obtained include property and violent crime rates in 2019 (City of Chicago Data Portal, 2019) and poverty rates (ACS, 2018). Some other demographic characteristics were also included in the analysis to provide a context for neighborhood environment, such as racial and ethnic compositions, percentage of no high school diploma, and percentage of children. Appendix eTable1 in the online supplement provides details regarding how these measures were created and from what sources the data used to create them were obtained. We also added individual locations of brownfield sites, parks, industrial corridors, rail yards, and major roads using a *turbopass* query of OpenStreetMap data (2022) in QGIS 3.22 software. Brownfield sites are empty lots or places previously developed, but not currently in use. These features highlighting neighborhood disadvantage are included to highlight dimensions of the physical environment that may intersect the experiences of PWID and have been shown to be associated with overdose risk (Tempalski et al., 2022) as well as downstream infections like HIV (Brawner et al., 2022).

### 2.3. Characterizing residential groups

We summarized demographic characteristics, risk behaviors, health outcomes, and social network measures for each residential group (i.e., urban, suburban, transient). For each measure included, we conducted bivariate analyses (Kruskal-Wallis rank sum test for continuous and ordinal variables, Pearson’s Chi-squared test for categorical variables with all expected cell counts >=5 or Fisher’s exact test for categorical variables with any expected cell counts < 5) to test for statistically significant differences between residential groups (see Table notes for more details). All statistical tests were 2-sided and significance levels are reported for each test. R version 4.0.2 was used to calculate statistical tests.

### 2.4. Point data visualization and kernel density estimate analysis

We explored the location of participant reported activity spaces to understand the distribution of risk activities across spaces. The Chicagoland metropolitan area includes the city of Chicago and its surrounding suburbs that span 16 counties in northeast Illinois, southeast Wisconsin, and northwest Indiana. Near suburbs include towns in Cook County surrounding Chicago and the bordering “collar counties,” namely DuPage, Kane, Lake, McHenry, and Will. The majority of activities occurred inside Cook and Collar counties (see Table 2 and Figure 2 in the Results section). Since our goal was to identify concentrated risk activity spaces (CRAS) by means of KDE, we restricted our sample to activities located exclusively within Cook and Collar counties. The reason for this is that far away activities (i.e., outliers) have considerable effects on KDE estimates because they artificially widen activity spaces, leading in turn to less concentrated area estimates that are less useful for intervention planning.

**Table 2.** Demographic characteristics by suburban, transient, and urban groups.

| Variable | All,<br>N = 258 | Suburban,<br>N = 72 | Transient,<br>N = 77 | Urban,<br>N = 109 | p-value |
| --- | --- | --- | --- | --- | --- |
| Race/Ethnicity |  |  |  |  | .025 |
| Hispanic | 65 (25%) | 23 (32%) | 15 (19%) | 27 (25%) |  |
| Non-Hispanic White | 153 (59%) | 45 (62%) | 50 (65%) | 58 (53%) |  |
| Other | 40 (16%) | 4 (6%) | 12 (16%) | 24 (22%) |  |
| Gender |  |  |  |  | .8 |
| Male | 185 (72%) | 49 (68%) | 56 (73%) | 80 (73%) |  |
| Female | 72 (28%) | 23 (32%) | 21 (27%) | 28 (26%) |  |
| Transgender | 1 (0.4%) | 0 (0%) | 0 (0%) | 1 (0.9%) |  |
| Age |  |  |  |  | .3 |
| 18-25 | 48 (19%) | 17 (24%) | 14 (18%) | 17 (16%) |  |
| 26-29 | 114 (44%) | 36 (50%) | 33 (43%) | 45 (41%) |  |
| 30-34 | 49 (19%) | 10 (14%) | 18 (23%) | 21 (19%) |  |
| 35 + | 47 (18%) | 9 (12%) | 12 (16%) | 26 (24%) |  |
| Region of birth |  |  |  |  | .003 |
| Chicago | 97 (38%) | 20 (28%) | 21 (27%) | 56 (51%) |  |
| Suburb, within IL | 113 (44%) | 38 (53%) | 38 (49%) | 37 (34%) |  |
| Outside IL | 48 (19%) | 14 (19%) | 18 (23%) | 16 (15%) |  |
| Homeless (Yes) | 167 (65%) | 21 (29%) | 66 (86%) | 80 (73%) | <.001 |
| Homeless (# of days) | 90 (30, 178) | 40 (20, 100) | 78 (30, 90) | 120 (60, 180) | <.001 |

**Note.** Median (IQR) was provided for continuous variables and frequency (percentage) was provided for binary variables. eTable 3 in Appendix details missing information for all variables. P-value was calculated with Kruskal-Wallis rank sum test for continuous variables, Pearson's Chi-squared test for categorical variables with all expected cell counts $\geq 5$ or Fisher's exact test for categorical variables with any expected cell counts $< 5$ . "Other" in Race/Ethnicity include Black/African American, mixed, and others.

Extensively discussed in Gramacki (2018), KDE constitutes a smoothing technique that approximates the distribution of point data along a continuous surface, so that one could understand how likely an event is to occur across spaces, or the “intensity” of activities. As a visualization tool, KDE has been used to reveal “hot regions” with a high density of occurrences (Bornmann & Waltman, 2010; Carlos et al., 2010). Additionally, such an approach has also been employed to identify individuals’ activity spaces based on daily activity locations and create environmental feature exposure measures in different research areas, such as those regarding foodscape (Crawford et al., 2014; Kestens et al., 2010) and the environment associated with weight-related behaviors and outcomes (Zenk et al., 2011).

In this paper, we produced KDE for the spatial distribution of different types of risk behaviors, including drug purchase, drug injection, and sex partner meetings, for each residential group of interest (i.e., urban, suburban, and transient individuals). All KDE analyses were performed using R, version 4.0.2 (R Foundation for Statistical Computing). We present the results with Gaussian smoothing, and the bandwidth is obtained by the Least Square Cross Validation (LSCV) schemes and ad-hoc criteria. Bandwidth was selected using data-driven techniques in part due to the lack of knowledge and published research of social-spatial interactions in this population. See Appendix for technical details and discussions regarding the decisions of kernel type and bandwidth.

The KDE analysis generates an estimate for the density of risk behavior locations. To further identify concentrated risk activity space (CRAS) for each residential group, we examine how the number of individuals involved increases as KDE contour range goes up (i.e., area gets larger). For instance, the 1% contour range of KDE identifies the area within which 1% of the activity locations occur, and we calculate how many individuals are involved in such activities. We continue such calculation: as the contour range increases from 1% to 100% more and more individuals in each residential group get involved in the corresponding range. Similar to the “elbow method” used to determine optimal number of clusters in clustering analysis, the concentrated risk activity space (CRAS) for each residential group is determined at the point where the number of individuals involved does not increase significantly with another expansion in contour range. The goal of identifying CRAS is to suggest future intervention avenues. Since a larger area may suggest more cost, the aim here is to identify a relatively small area (or areas) with more individuals involved.

### 2.5. Characterizing the concentrated risk activity spaces

To further contextualize the CRAS for each residential group (urban, suburban, and transient), we identified the census tracts comprising each CRAS to examine multiple measures of neighborhood environment (see Measures section). We also conducted bivariate analyses for each neighborhood measure to test for statistically significant differences between the identified CRASs and other areas in Chicago. All statistical tests were 2-sided, and we reported significance levels for each test. R version 4.0.2 was used for all statistical analyses.

### 2.6. Spatial visualizations

We developed multiple maps to inspect, explore, and present data and findings. We first developed a choropleth map of participants by census tract, including county and Chicago boundaries and a reference base map for context. We next mapped activity places across the full extent of the original geographic survey, both individually and all together. These maps helped informed which areas to include, and exclude, for next steps of the analysis. We mapped KDEs of the final study area, stratified by activity place type and residential group. Finally, we mapped the Chicago-specific locations of CRAS, including a zoomed in view of overlapping residential group CRASs by Census tracts that also include locations of key built environment features such as parks and industrial corridors. All mapping was done in QGIS 3.22, with the exception of KDE plots that were visualized in R.

## 3. Results

### 3.1. Residential groups

Among 258 participants, 72 (28%) were identified as suburban, 77 (30%) as transient, and 109 (42%) as urban based on their reported places of residence during the last year (see Measures section). Figure 3 provides a contextual map showing spatial distribution of all reported residential addresses in the past year.

**Figure 3.**
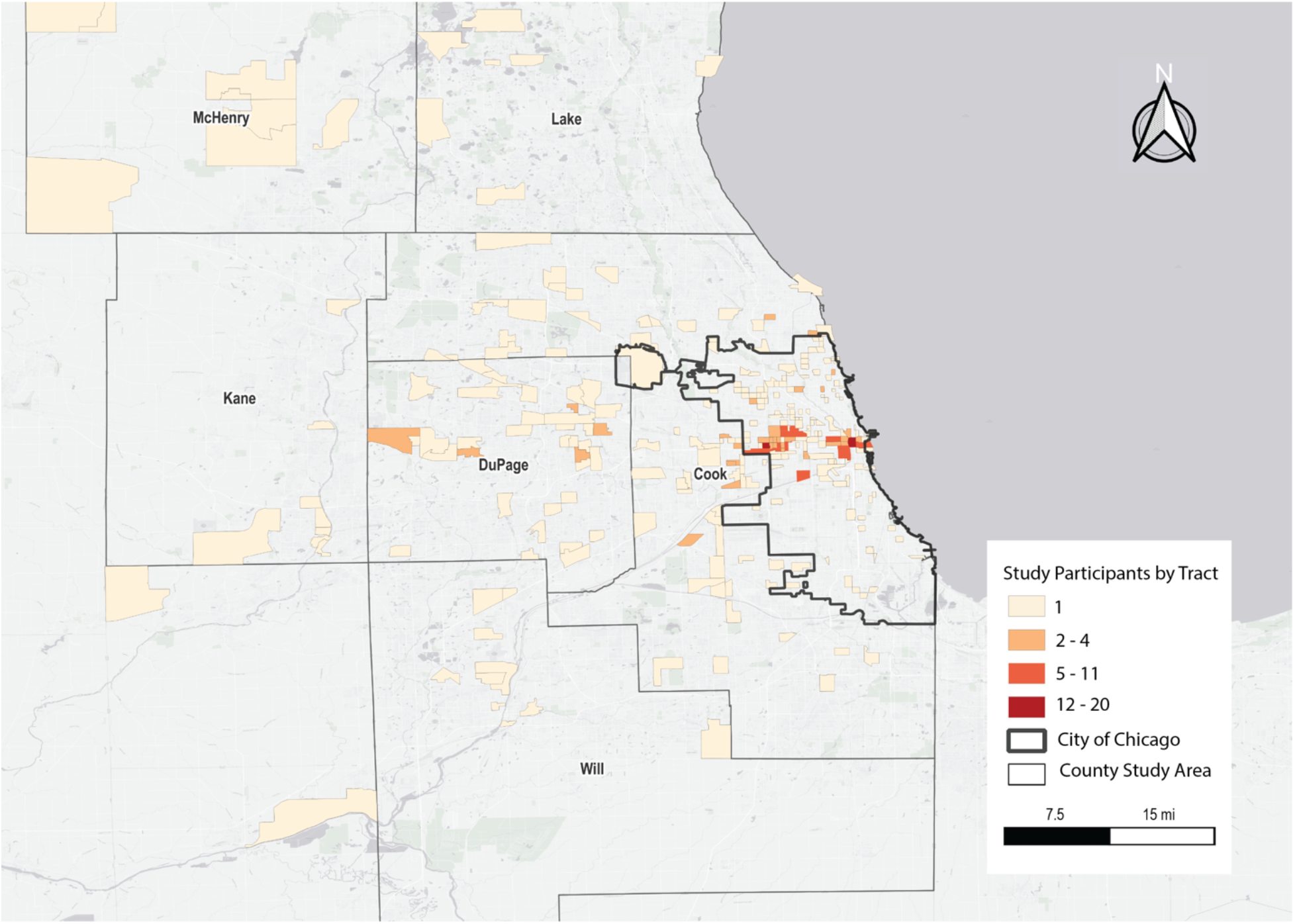
Spatial distribution of participants: residential addresses are aggregated by census tract and visualized as a thematic map, highlighting City of Chicago and study area county boundaries.

### 3.2. Sociodemographic variation

We stratified demographic characteristics, risk behaviors, health outcomes, and injection network measures by each residential group (Table 2–4). We first reported whether and how the three residential groups vary in these measures. In terms of demographic backgrounds, all individuals who identified themselves as non-Hispanic Black were either transient or urban, meaning at least one of their places of residences in the last year was within Chicago. The majority (94%) of suburban individuals were either self-identified as Hispanic or non-Hispanic white. There were no statistically significant differences across the three residential groups (i.e., urban, suburban, transient) in terms of age or gender. 72% of all individuals were male and 63% of them were young PWID aged between 18 and 29. The urban group had the largest proportion of PWID born in Chicago (51%), compared to 28% in the suburban group and 27% in the transient group. Importantly, more transient (86%) and urban (73%) than suburban (29%) PWID reported experiencing homelessness in the past 6 months, with the urban group reported the largest number of days experiencing homelessness (median [IQR]: 120 [60, 180]), followed by the transient group (median [IQR]: 78 [30, 90]) and suburban group (median [IQR]: 40 [20, 100]).

### 3.3. Risk behaviors and health outcomes

Overall, more PWID in the transient and urban groups reported significantly higher risk behaviors compared to the suburban group, including injecting crack, speedball, or methamphetamine; and backloading (i.e., shooting up with a syringe after someone else has squirted drugs into in from their needle) (Table 3). Notably, PWID in the transient group reported the largest prevalence of cooker sharing (84%), followed by the suburban (70%) and urban groups (63%). In terms of health outcomes, significantly more urban PWID (45%) tested positive for HCV, compared to 29% in the transient group and 13% in the suburban group. Only three PWID tested HIV positive, and they were either in the transient or urban group. Though not statistically significant at the 0.05 level (p = 0.06), the transient group reported the smallest number of months since last overdose (median [IQR]: 4 [2, 19]), followed by the urban group (median [IQR]: 7 [2, 26]) and suburban group (median [IQR]: 12 [3, 28]).

**Table 3.** Risk behaviors and health outcomes by suburban, transient, and urban groups.

| Variable | All,<br>N = 258 | Suburban,<br>N = 72 | Transient,<br>N = 77 | Urban,<br>N = 109 | p-value |
| --- | --- | --- | --- | --- | --- |
| Used crack | 215 (84%) | 58 (82%) | 69 (90%) | 88 (81%) | .2 |
| Injected crack | 101 (39%) | 20 (28%) | 36 (47%) | 45 (41%) | .050 |
| Injected speedball | 112 (43%) | 24 (33%) | 35 (45%) | 53 (49%) | .12 |
| Used methamphetamine | 72 (28%) | 14 (20%) | 25 (32%) | 33 (30%) | .2 |
| Injected<br>methamphetamine | 59 (23%) | 8 (11%) | 25 (32%) | 26 (24%) | .008 |
| Receptive syringe sharing | 106 (42%) | 30 (42%) | 36 (47%) | 40 (38%) | .5 |
| Sharing cookers | 179 (72%) | 50 (70%) | 63 (84%) | 66 (63%) | .011 |
| Backloading | 95 (38%) | 19 (27%) | 40 (53%) | 36 (35%) | .004 |
| Experienced overdose past<br>6 months | 93 (36%) | 21 (29%) | 36 (47%) | 36 (33%) | .057 |
| Revived with Naloxone<br>past 6 months | 81 (31%) | 17 (24%) | 31 (40%) | 33 (30%) | .086 |
| Months since last<br>overdose | 7 (2, 25) | 12 (3, 28) | 4 (2, 19) | 7 (2, 26) | .060 |
| Months since last<br>Naloxone | 8 (2, 27) | 12 (5, 27) | 5 (2, 25) | 6 (2, 27) | .14 |
| HCV positive | 77 (32%) | 9 (13%) | 22 (29%) | 46 (45%) | <.001 |
| HIV positive | 3 (1.2%) | 0 (0%) | 2 (2.7%) | 1 (1.0%) | .5 |

**Note.** Median (IQR) was provided for continuous variables and frequency (percentage) was provided for binary variables. eTable 4 in Appendix details missing information for all variables. P-value was calculated with Kruskal-Wallis rank sum test for continuous variables, Pearson's Chi-squared test for categorical variables with all expected cell counts $\geq 5$ or Fisher's exact test for categorical variables with any expected cell counts $< 5$ .

### 3.3. Social networks

As shown in Table 4, the suburban, transient, and urban groups also demonstrated substantial differences in terms of their egocentric injection network measures. First, the transient group had the largest injection networks with the most non-redundant ties (median [IQR] network degree 4 [3,6]; network effective size 2.71 [1.84, 4.00], *p* = 0.002 for both measures). Consistently, the transient group also had the smallest tie density (median [IQR]: 0.86 [0.69, 1.00], *p* = 0.014). PWID in the suburban group had the most alters living outside of Cook County (50% [0%, 100%], *p* < 0.001). These alters were also younger and more similar in age than alters of PWID in the urban and transient group (mean age of alters is 33 [30, 37] for urban, 33 [30, 36] for transient and 31 [27, 35] for suburban, *p* = 0.004; maximum alter age is 40 [32, 50] for urban, 40 [35, 48] for transient and 34 [29, 40] for suburban, *p* < 0.001; age standard deviation of alters is 6.1 [4.3, 9.1] for urban, 7.1 [4.6, 9.3] for transient and 3.6 [1.5, 6.5] for suburban, p < 0.001). Sexual and support network measures showed similar patterns with a few unique features (see eTable 5 in the Appendix). For instance, compared to urban and transient groups, the suburban PWID reported stronger connections with their sexual and support network members, but there were no significant differences in terms of network density across three groups.

**Table 4.** Injection network measures by suburban, transient, and urban groups.

| Variable | All,<br>N = 258 | Suburban,<br>N = 72 | Transient,<br>N = 77 | Urban,<br>N = 109 | p-value |
| --- | --- | --- | --- | --- | --- |
| Network Degree | 3.00<br>(2.00, 5.00) | 3.00<br>(2.00, 4.00) | 4.00<br>(3.00, 6.00) | 3.00<br>(2.00, 5.00) | .002 |
| Mean strength of ties | 3.00<br>(2.75, 3.60) | 3.00<br>(2.84, 3.62) | 3.00<br>(2.71, 3.33) | 3.00<br>(2.66, 4.00) | .5 |
| Age standard deviation of alters | 5.9 (3.5, 8.7) | 3.6 (1.5, 6.5) | 7.1 (4.6, 9.3) | 6.1 (4.3, 9.1) | <.001 |
| Average age of alters | 32 (29, 36) | 31 (27, 35) | 33(30, 36) | 33 (30, 37) | .004 |
| Max age of alters | 38 (32, 47) | 34 (29, 40) | 40 (35, 48) | 40 (32, 50) | <.001 |
| % alters living in Cook county | 100<br>(50, 100) | 50<br>(0, 100) | 100<br>(60, 100) | 100<br>(97, 100) | <.001 |
| Residence homophily | 100<br>(50, 100) | 50<br>(0, 100) | 100<br>(50, 100) | 100<br>(85, 100) | <.001 |
| Network effective size | 2.06<br>(1.25, 3.05) | 1.75<br>(1.32, 2.48) | 2.71<br>(1.84, 4.00) | 2.00<br>(1.00, 2.93) | .002 |
| Network tie density | 0.93<br>(0.73, 1.00) | 1.00<br>(0.83, 1.00) | 0.86<br>(0.69, 1.00) | 0.97<br>(0.80, 1.00) | .014 |

**Note.** Median (IQR) was provided for each variable. eTable 5 in Appendix details missing information for all variables. P-value was calculated with Kruskal-Wallis rank sum test.

**Table 5.**
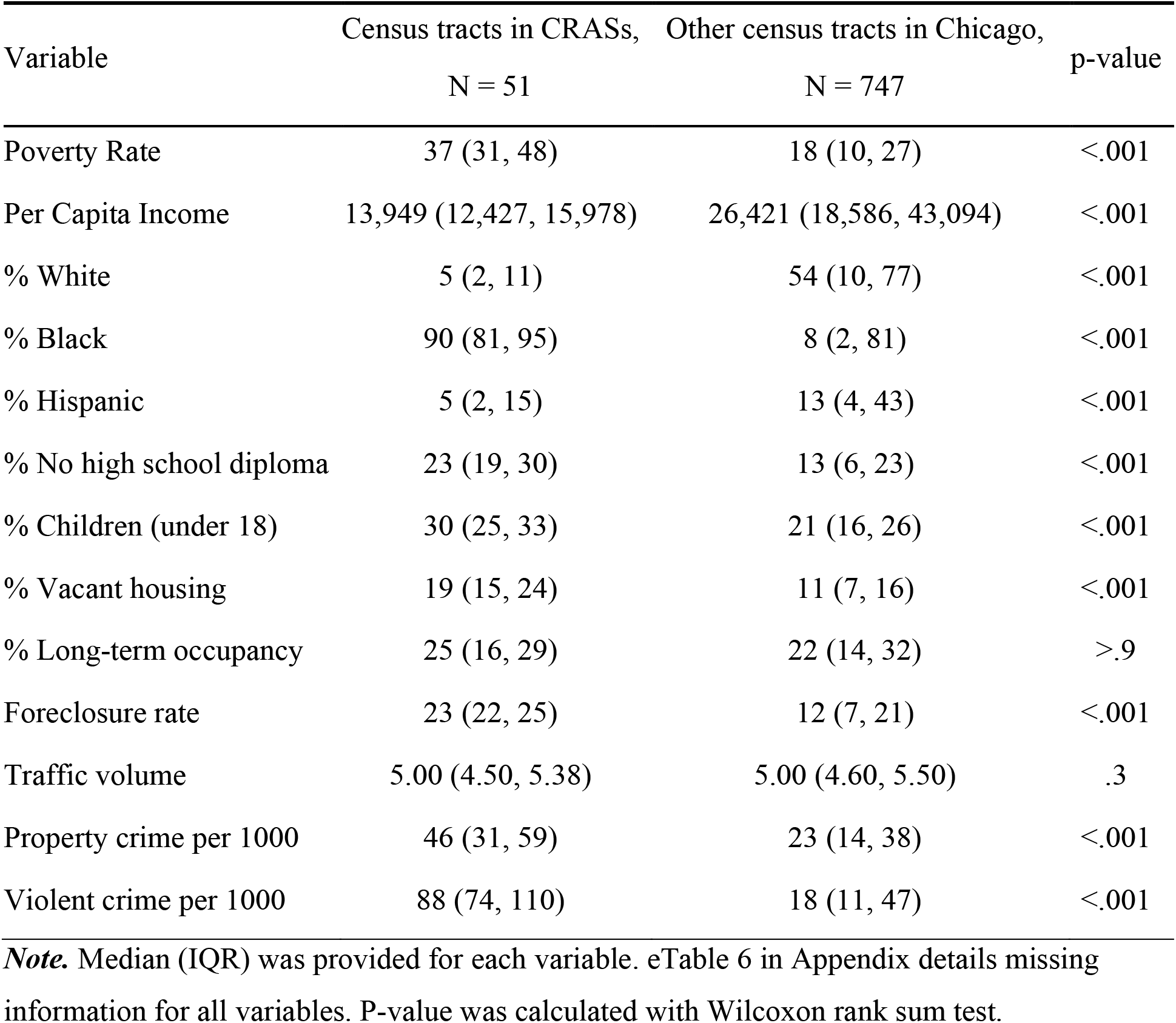
Neighborhood characteristics of identified concentrated risk activity spaces

| Variable | Census tracts in CRASs, | Other census tracts in Chicago, | p-value |
| --- | --- | --- | --- |
|  | N = 51 | N = 747 |  |
| Poverty Rate | 37 (31, 48) | 18 (10, 27) | <.001 |
| Per Capita Income | 13,949 (12,427, 15,978) | 26,421 (18,586, 43,094) | <.001 |
| % White | 5 (2, 11) | 54 (10, 77) | <.001 |
| % Black | 90 (81, 95) | 8 (2, 81) | <.001 |
| % Hispanic | 5 (2, 15) | 13 (4, 43) | <.001 |
| % No high school diploma | 23 (19, 30) | 13 (6, 23) | <.001 |
| % Children (under 18) | 30 (25, 33) | 21 (16, 26) | <.001 |
| % Vacant housing | 19 (15, 24) | 11 (7, 16) | <.001 |
| % Long-term occupancy | 25 (16, 29) | 22 (14, 32) | >.9 |
| Foreclosure rate | 23 (22, 25) | 12 (7, 21) | <.001 |
| Traffic volume | 5.00 (4.50, 5.38) | 5.00 (4.60, 5.50) | .3 |
| Property crime per 1000 | 46 (31, 59) | 23 (14, 38) | <.001 |
| Violent crime per 1000 | 88 (74, 110) | 18 (11, 47) | <.001 |

**Note.** Median (IQR) was provided for each variable. eTable 6 in Appendix details missing information for all variables. P-value was calculated with Wilcoxon rank sum test.

### 3.5. Risk activity spaces

Figure 4 shows the locations of spaces of all risk activities for the suburban, transient, and urban groups. Most activities took place within Cook and Collar counties (see Appendix for summary statistics). In general, the urban group reported the fewest risk activities outside the Cook and Collar counties’ geographic boundaries, while the transient group reported the most outside these boundaries (though this was still a very small proportion all risk activities). Compared to sex partner meeting locations, injection locations and drug purchasing locations were more concentrated within Cook and Collar counties. In fact, the urban group reported purchasing and injecting drugs only within Cook and Collar counties’ geographic boundaries. Given such patterns across groups and different activities, we also calculated how far each PWID traveled for different activities (Figure 3 and eTable 7). Consistent with the point data visualization, the urban group had the shortest travel distance among the three residential groups, for all three types of risk activities (drug purchasing 3.62 [1.38, 5.12], drug injection 2.44 [1.01, 4.05], or meeting sex partners 3.16 [0.96, 6.74]) (Figure 3 and last column in eTable 7). Meanwhile, the suburban group traveled the furthest for drug purchasing (median [IQR]: 13.51 [5.94, 21.34]), but traveled less than the transient group for drug injection and meeting sex partners. For example, the median travel distance for meeting sex partner in the suburban group is 6.93 miles (IQR, [2.99, 17.51]), while the median distance in the transient group is 13.43 miles (IQR, [8.02, 51.24]). All comparisons among three residential groups are statistically significant with p < 0.001.

**Figure 4.**
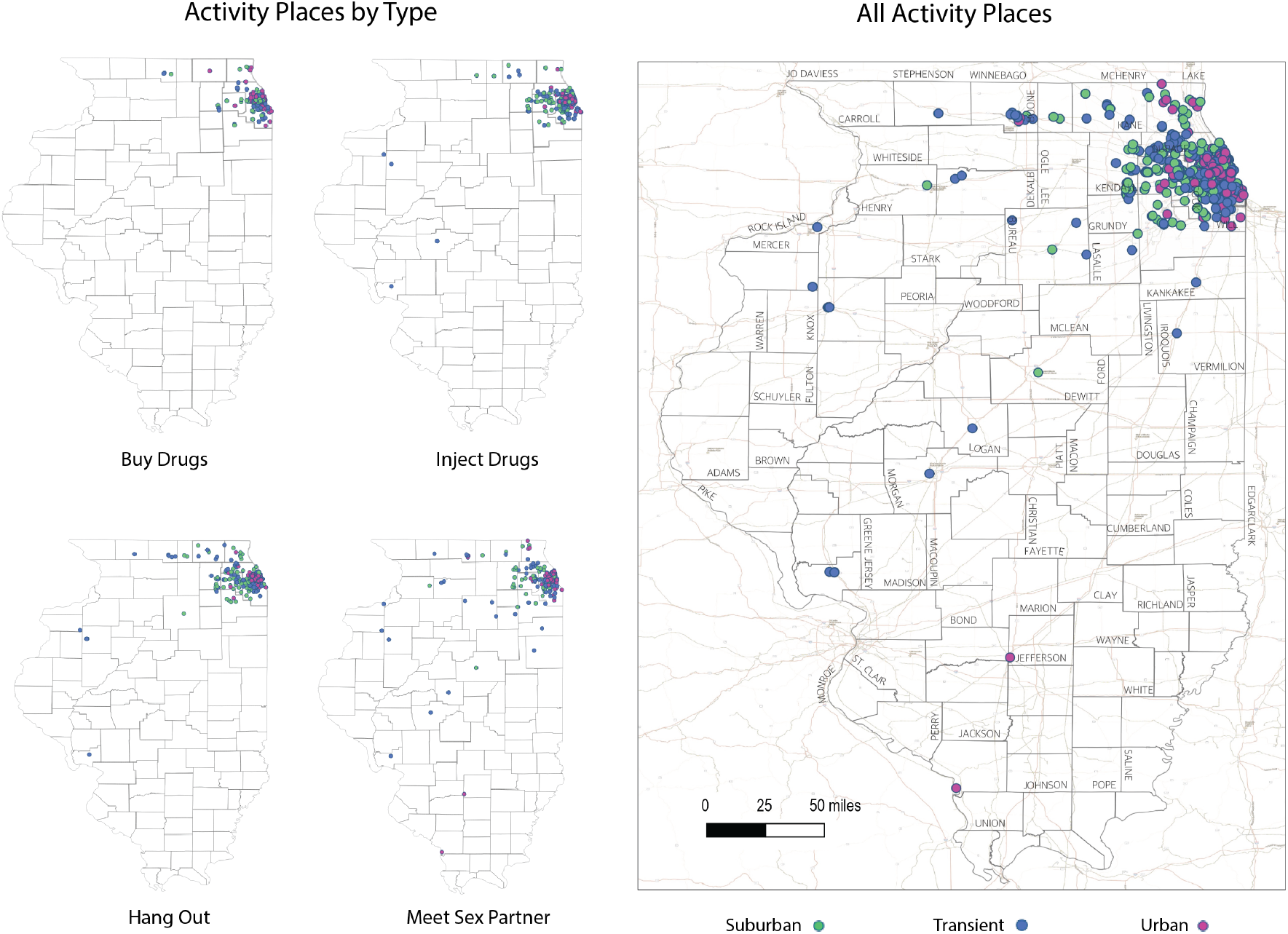
Spatial distribution of all activity spaces in Illinois, by activity topic and participants’ residential group (suburban, transient, urban).

**Figure 5.**
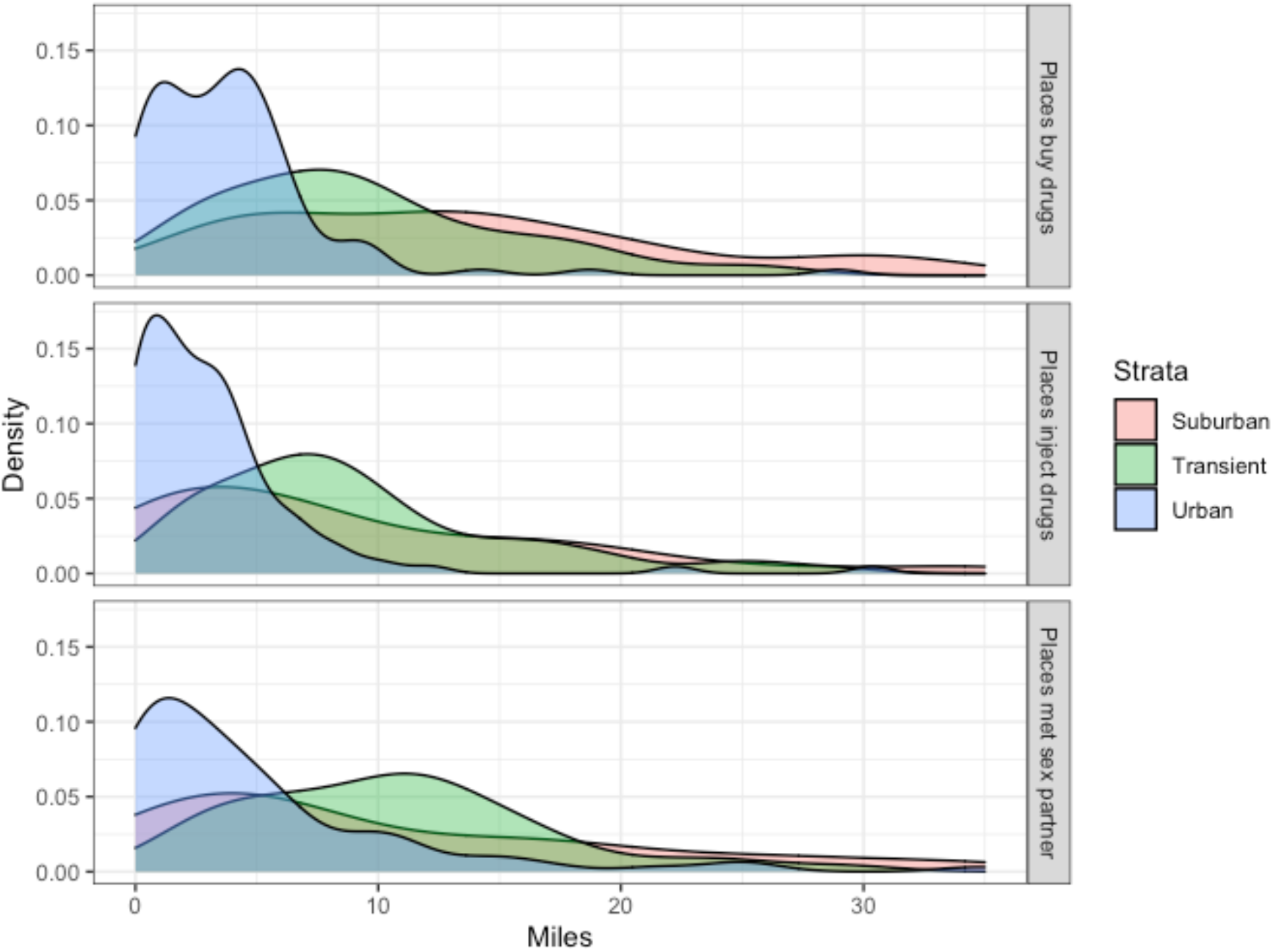
Average travel distance (miles) by activity topic and participants’ residential group (suburban, transient, urban). ***Note***. The distance metrics are calculated for those PWID who reported at least one geographic location data for the particular activity. Specifically, travel distance to meeting sex partners is reported for 82.6% participants (n = 213) who reported at least one location for meeting sex partners. Distance to drug purchasing is reported for 98.4% participants (n = 254) and distance to drug injection is reported for 97.3% participants (n = 251).

In Figure 6, we presented the KDE results for each risk activity by residential group. First, for activities of drug injection and meeting sex partners, the suburban group had the most dispersed locations while the urban group had the most concentrated locations in a smaller area. Additionally, consistent with the point data visualization in Figure 4, most of the drug purchases occurred in the same spot along the west side of Chicago regardless of the residential group, suggesting an impactful area for possible public health interventions. In contrast, places meeting sex partners were more widely distributed across geographic spaces. In fact, the KDE results (Figure 6C) demonstrate that even the urban group (which primarily purchased and injected drugs within Chicago) also met sex partners outside of Chicago (even a few outside Cook and Collar Counties, see Figure 4).

**Figure 6.**
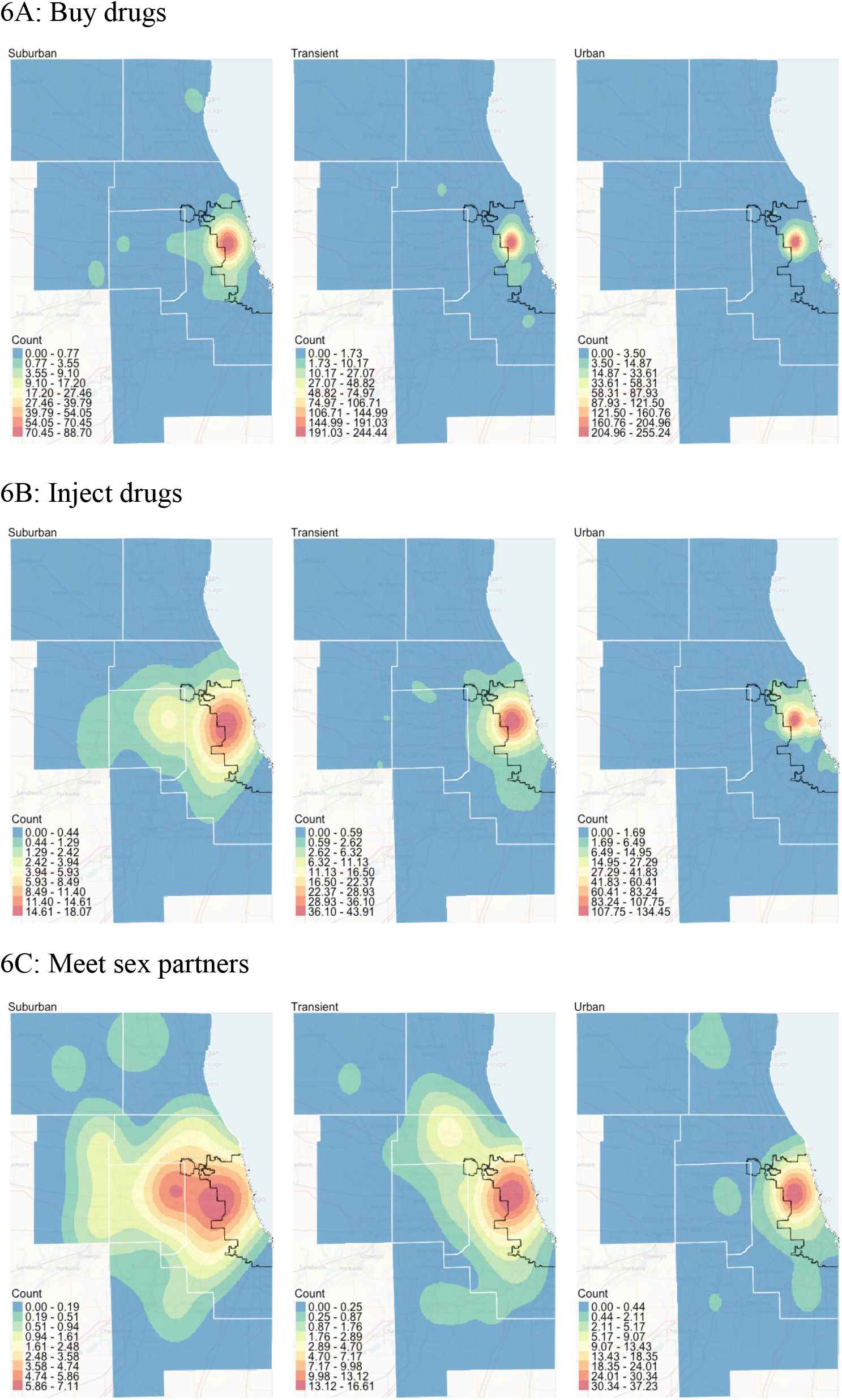
Kernel density estimates (KDE) by activity topic and residential group (suburban, transient, urban).

Based on the KDE results, we further identified concentrated risk activity spaces (CRAS) for each residential group. As shown in Figure 7, we plotted out how the number of individuals included changed against KDE contour ranges, from which we observed that most individuals’ risk activities are concentrated within a limited range. For example, the 7% KDE contour covers risk activities of 87 (out of 109) individuals from the urban group. Similarly, the 8% KDE contour covers risk activities of 63 (out of 70) individuals from the suburban group; and the 9% KDE contour covers risk activities of 72 (out of 76) individuals from the transient group. The results remained almost the same if we excluded sex partner meeting locations to consider only drug purchasing and injection locations as risk activities.

**Figure 7.**
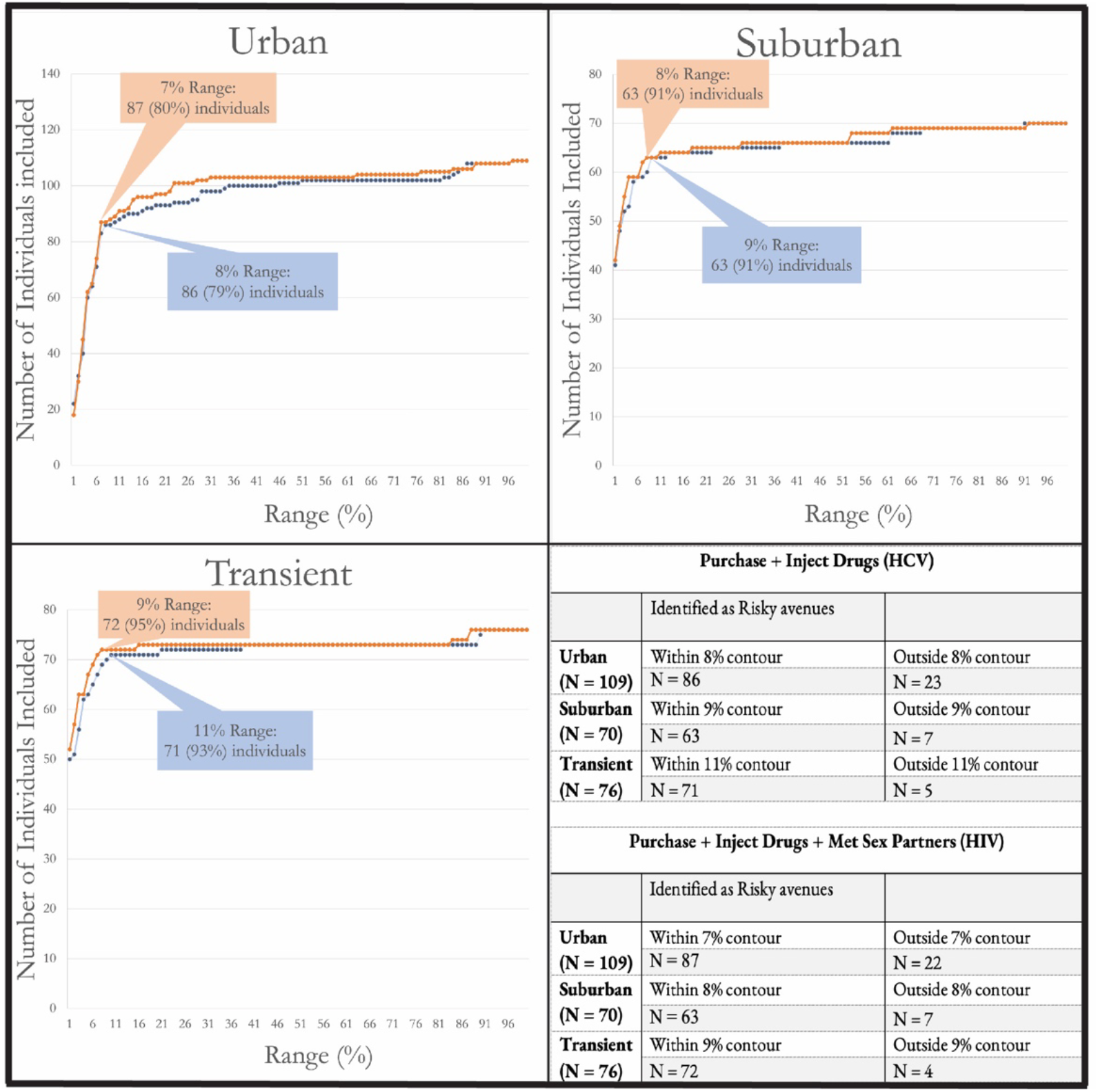
Identify concentrated risk activity spaces (CRASs) based on KDE by participants’ residential group (suburban, transient, urban).

To further contextualize the CRAS areas, we identified the census tracts linked with the KDE contours for each residential group (urban, suburban, and transient). As shown in Figure 8, the urban group had the most compact CRAS, while the suburban group had the largest one. Importantly, the three CRASs are geographically overlapped: the urban CRAS was nested within the transient CRAS, and the transient CRAS was nested within the suburban CRAS. In the same figure, built environment features are highlighted prominently in the overlapping CRAS areas. The urban CRAS, which included core activity spaces for all residential groups, is dominated by brownfield sites, industrial buildings, rail corridors, and a large park. The CRAS are bounded along the I-290 on the Southern end of the map, sharing a public transit rail line.

**Figure 8.**
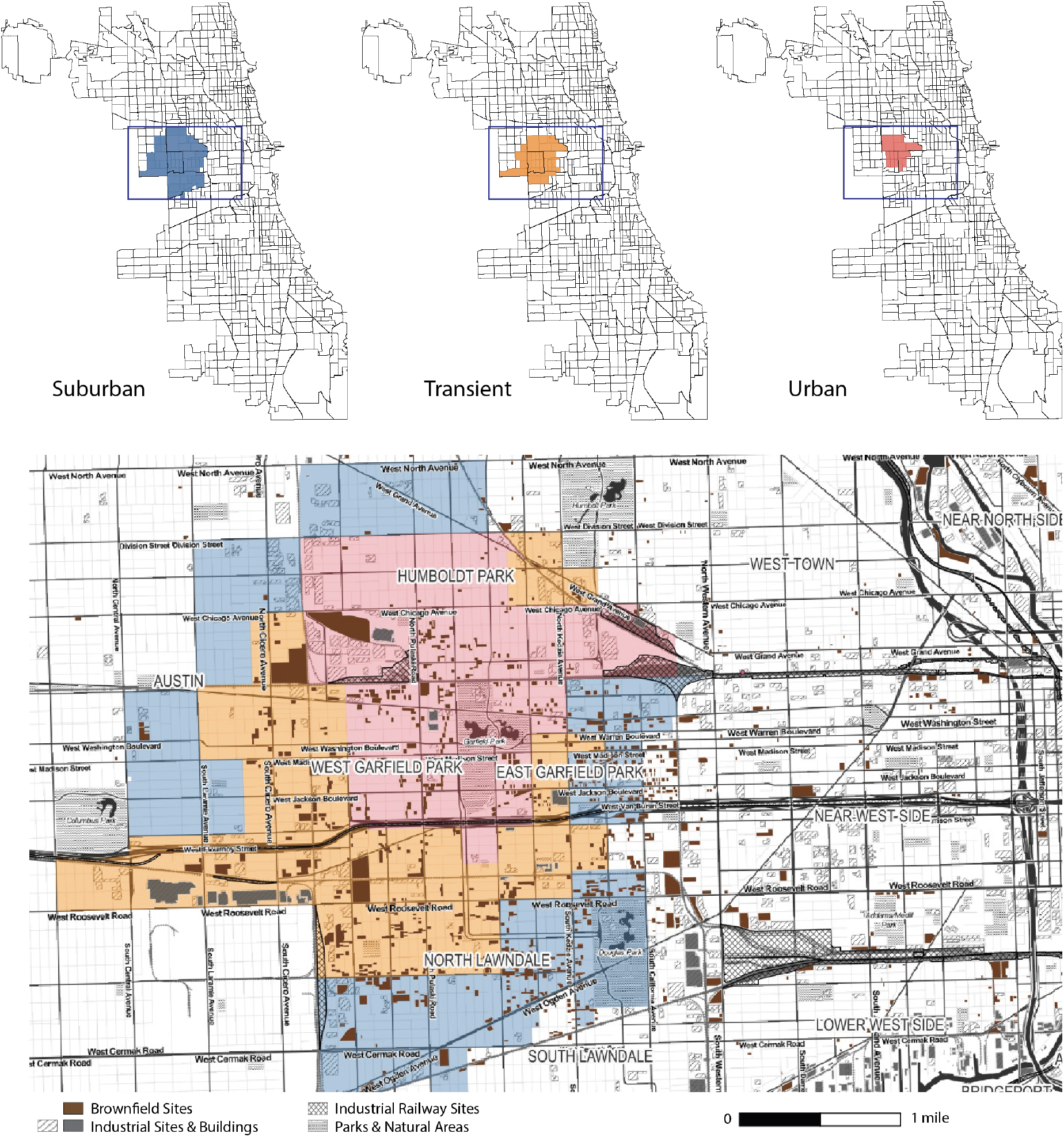
Concentrated risk activity spaces (CRASs) for all participant groups (urban, suburban, and transient).

We used the Census tracts comprising CRASs to examine multiple measures of neighborhood disorder and disadvantage. In Table 5, we summarized neighborhood measures for 51 Census tracts linked to identified CRASs for all three residential groups taken together (first column). Compared to the other census tracts in Chicago (second column), the identified CRASs had significantly higher poverty rates, lower income rates, higher percentage of adult residents without a high school diploma, more children under 18, higher vacancy rates, higher foreclosure rates, and both higher property and violent crime rates (all with p-value < 0.001). The percentage of long-term occupancy in CRAS was not significantly different from other areas.

## 4. Discussion

Our study reports on geographic variation within a large, connected region in injection behaviors and social network features to inform risk mitigation and intervention development for young PWID. Despite having an expansive, multi-county geographic extent of PWID showing heterogeneity in mobility, traveling behaviors and social network structures, 93% of study participants had IDU interactions within a singular, centralized geographic space (CRAS) identified from our spatial analysis, suggesting venues for place-based intervention. Notably, this identified CRAS area also overlaps with a recent study’s reported geographical clustering concentration of fentanyl-involved overdose deaths in Cook County from 2014 to 2018 (Nesoff et al., 2020). Below we discuss key findings following the multilevel structure in the socio-ecological model, namely individual, social interactions at the interpersonal level, and community level. Notably, this organization is also consistent with our research design: we identified suburban, urban, and transient PWID groups based on their recent residential addresses and then considered the potential heterogeneity in terms of social network characteristics and locations of risk behaviors. Throughout, we also discuss venues for future research and insights for interventions to address the syndemic.

At the individual level, the young PWID in this study are predominantly born in and/or reside in the surrounding suburban areas. This is consistent with our prior research studies in Illinois and the pattern observed in attendants of a large multisite and mobile-van based SSP (Boodram et al., 2010, 2015; Mackesy-Amiti, 2021; Mackesy-Amiti et al., 2012; Thorpe et al., 2001). Inspired by activity space literature, our grouping of suburban, urban, and transient PWID based on their recent residential addresses led to a few important insights regarding mobility and travel behaviors. For example, in this study, 72% of young PWID in the suburban residential group were also born in suburban areas, with a similar proportion for those in the transient (73%) and almost half (49%) of the urban residential groups. Here, as with many states, HIV and HCV prevalence in urban areas are dramatically higher than their surrounding suburbs (Chicago Department of Public Health, 2010; Cook County Department of Public Health, 2010). Interaction in risk activity spaces common to both urban and suburban PWID (e.g., drug market areas) may be key to understanding the evolution of the increasing HIV and HCV prevalence in suburban areas.

Additionally, we identified distinct travel behaviors within each PWID residential group. Overall, we found that young suburban PWID traveled farther spatially than transient or urban PWID to purchase drugs while transient— and to a lesser extent suburban— PWID traveled farther than the urban PWID to inject drugs and meet sex partners. When considering the distance traveled to risk activity locations, our study provides new insights for the important question of “how far are persons willing to travel?” Such question is of great importance as it helps understand underlying drivers of traveling behaviors and optimize resource allocations and placement. For example, urban dwellers peaked in travel to an activity within about one mile to inject drugs and didn’t go beyond ten; suburban dwellers peaked around three miles, but some would travel more than twenty miles for the same activity. Furthermore, distance traveled varied according to activity. Engagement in meeting sex partners was fairly spatially expansive for all three residential groups, with some traveling to distant areas from their residences to meet sex partners. KDE results likewise suggested that the CRAS for suburban PWID was more spatially expansive than those of the other two groups, and that the CRAS for urban PWID was the most spatially compact. These patterns could in part be explained by variation in transportation methods and access across the three residential groups. Suburban and transient PWID may be more likely to have access to a vehicle, whereas urban PWID may be more likely to utilize public transit, which could make further travel more inconvenient and less desirable. Such a pattern is also consistent with the theoretical assertion by Rhodes et al. (2006) that access to public transportation and/or to main thoroughfares and highways influences PWID’s injection activity.

Moreover, our study was consistent with extant literature on PWID of all ages that show residential transience and similar measures of housing instability, which are also associated with higher likelihood of a number of injection- and substance use-related harms among PWID (Arum et al., 2021; Boodram et al., 2015, 2018; German et al., 2007). However, in our sample of young PWID, the residential transience was not associated with HCV antibody status; indeed, urban young PWID were significantly more likely to be HCV-positive than their counterparts, which can be partially explained by two factors. First, much of the extant literature has defined transience as having a specific minimum number of residences (anywhere) within a specific time period, while the present study is instead capturing a more specific type of transience that Boodram et al. (2015) referred to as “crossover” transience – i.e., having residences in both urban and suburban areas within a connected region in a given period of time. Such conceptualization is key to understanding HIV and HCV transmission and informing network-based interventions in large metropolitan areas like Chicago and others (e.g., Baltimore) with varying levels of mobility among PWID who might serve a ‘bridges’ between high (e.g., urban) and low (e.g., suburban) encompassing regions (Boodram et al., 2010). Second, our sample is generally younger (88% were ages 18-34, with urban having largest group ages 35 and older), with corresponding shorter time injecting and fewer resulting transmission opportunities, compared to samples from the extant literature.

At the interpersonal level, we also find evidence that suburban, urban, and transient PWIDs have different network characteristics. Specifically, we found that transient PWID had the largest and least dense injection networks. This population represents those who had lived both inside and outside of Chicago in the last year and, therefore, are likely to interact and bridge/connect people in multiple, distant spaces. At the same time, travel cost, access to transportation, and limited resources may limit mobility; yet our findings suggest that this transient group of PWID remains highly mobile. Suburban PWID had the most network members living outside of Cook County, which is intuitive since the suburbs are closer to the edges of Cook County and had network members who were significantly younger relative to the network members of transient or urban PWID.

At the community level, a key finding of our analyses is that the CRAS for each of the three residential groups completely overlapped, with the CRAS for urban PWID nested within the CRAS for transient PWID, and with the CRAS for transient PWID nested within the CRAS for suburban PWID. This distinct spatial structure was persistent across different residential groups, despite varying travel behaviors and underlying factors of risk. A similar phenomenon has been characterized as “convection mixing” by Gesink et al 2020’s case study of the social geography of sexually transmitted infections (STI) within a vulnerable population. In their analysis, participants described a cyclical pattern of movement between suburban residences and urban destinations; assortative urban mixing nested within disassortative suburban participant mixing. This process was hypothesized to contribute to the persistence of STI transmission across core and distal outbreak areas (Gesink et al., 2020). The presence of convection mixing in our analysis has important implications for public health practice and intervention development to provide harm reduction and treatment services to PWID in Chicago to reduce risk of overdose and downstream infection transmissions. Specifically, it suggests that there are centralized geographically overlapped areas of risk activity that are relevant to and utilized by not only PWID living within Chicago but also PWID living within surrounding suburbs. Our geographic identification of one such centralized region, where PWID risk activity is concentrated, can be of use to public health practitioners and other interventionists who aim to maximize the benefit of limited resources towards providing needed services to local (and slightly less local – i.e., suburban) PWID.

Finally, the core activity place where “convection mixing” occurs across all residential groups exists in a place of great neighborhood disadvantage. CRASs for all residential groups of PWID were found to be significantly higher than other Chicago census tracts on a number of indicators of neighborhood social disorder, or key metrics of disadvantage. This is consistent with the extant literature that has found that most of these indicators (i.e., poverty rates, percentage of adult residents without a high school diploma, vacancy rates, foreclosure rates, property rates, and violent crime rates) are associated with greater substance use (Boardman et al., 2001; Cooper et al., 2016) and/or with risk for HCV, overdose, and other harms (Cerdá et al., 2013; Chen et al., 2022; Gu, 2021; Hembree et al., 2005). The presence of geographically concentrated brownfield sites in particular highlights the legacy of dis-investment in the area; once developed places have since been removed and are now grown over as empty lots nearby remote industrial corridors and isolated railway areas. Indeed, the legacy of Chicago’s West Side is historically documented as one of complex change, racial inequity, and a fight for resources over the past century (Seligman, 2005). Understanding the characteristics of PWID’s CRASs has important implications for intervention development and public health practice, as profiles of these characteristics could potentially be created for settings in order to predict risk for engagement in injection and drug purchasing behaviors, and to allocate resources and target interventions to those with highest predicted risk. Furthermore, our study findings support Brawner’s concept of “geobehavioral vulnerability” that underscore not just individual behaviors but both social and spatial interactions driving disease burdens (2014), also found in a recent Chicago-case studies of HIV social-spatial networks (Chen et al., 2019; Kolak et al., 2021). While these studies focused on HIV incidence, risk behaviors in PWID are relevant because of the implications for adverse outcomes in the individual, network, and downstream infections. Our study findings likewise support conceptual networks that integrated an intersectional view of drug related harm that consider multi-level risks across varying dimensions of the social-spatial environment (Collins et al., 2019; Rhodes, 2002) as well as recent extensions that incorporate the socio-built environment aspects directly (Tempalski et al., 2022). Future research may further explore how events of drug injection and meeting sex partners, also known as venues in social network literature (Frank et al., 2013; Menchik, 2019), interacts with neighborhood characteristics, with implications for PWID’s injection career and health.

### 4.1. Limitations

The present analyses are cross-sectional, exploratory, and largely descriptive. Causal relationships between geographic location and the other constructs of interest can in no way be inferred. Additionally, given the exploratory nature of this study, we did not attempt to examine the potentially complex interrelationships among all constructs of interest that would be supported by extant theory. Future research should employ multivariate analyses to examine the complex mechanisms through which multidimensional, interconnected factors shape PWID’s experience of the syndemic. Another limitation of the present study is that the COVID-19 pandemic began during the course of data collection. A number of constructs we measured may have been affected by the various social, economic, logistic, and psychological changes that the pandemic engendered for many people. Our future analyses of longitudinal data from this study will examine change over time in order to unpack the influence of the pandemic (and/or of other major events affecting the general population) on the spatial distribution of risk activity and social network characteristics among this sample of young PWID. We also acknowledge that the findings of the present study are not generalizable to populations of PWID outside of the sample studied, populations of PWID in other geographic regions, or those who are older or are of different racial/ethnic or socio-linguistic backgrounds. The population studied was not originally sampled using a spatially random approach, but rather recruited through a specific process that may result in spatially concentrated communities. At the same time, while the original population studied resides in a geography expansive area across multiple counties in Northeastern Illinois, the vast majority engaged in specific activities in a specific location of overlapping geographic locations. An exploratory, hypothesis-generating analysis can thus still derive meaning from social-spatial behavioral patterns emergent in smaller population samples that may be difficult to survey with spatially random designs. Additionally, we attempt to characterize risk activity spaces with self-reported place locations by survey participants. We did not have access to complete movement of the true activity space geographic extent of each individual. To maintain privacy of individuals in a vulnerable population, snapshots of activity spaces crucial to the understanding of geo-behavioral vulnerability were instead included. This additional aspect, which can be identified using GPS-data, is common in activity space literature and may be an area of future research.

## 5. Conclusions

Our study represents an important exploratory step in examining geographic variation both in PWID’s risk activity spaces (i.e., injecting drugs, purchasing drugs, and meeting sex partners) and in the characteristics of PWID’s social networks. We found that suburban PWID travel more extensively to engage in risk activities than do urban PWID, and that PWID’s engagement in drug injection and drug purchasing concentrates geographically into spaces (CRASs) that overlap across residential groups (urban, suburban, and transient) of PWID. This finding can be leveraged into maximizing resources and efforts to implement harm reduction interventions in centralized CRAS locations that will be able to serve PWID who live in a somewhat expansive surrounding area (i.e., both PWID living in Chicago and PWID living in its surrounding suburbs). We also found that CRASs across residential groups of PWID are higher on a number of location and space level indicators of social disorder and social disadvantage than other Chicago census tracts, suggesting that future studies can develop profiles of risk for specific settings based on such characteristics (for which data are readily available), in order to further improve allocation of resources for substance use prevention and harm reduction.

While our findings are not directly generalizable to PWID in other large metropolitan areas, the perspective and analytical approach utilized to integrate both risk activity spaces and social networks have important implications for future studies. Such approaches could be especially useful for areas with sub-groups (e.g., transient populations) and activity spaces (e.g., outdoor drug markets) that could potentially serve as bridges between areas of high and low HIV and HCV prevalence that could be targeted for intervention development and policy implementation.

## Data Availability

The data that support findings of this study are available upon reasonable request to the authors.

## Acknowledgement

This work was supported by the National Institutes on Drug Abuse (NIDA) [R01DA043484].

