## Supplemental for "Social-spatial network structures among young urban and suburban persons who inject drugs in a large metropolitan area"

### Appendix (online supplemental materials)

**eTable 1.** Neighborhood measure definitions and data sources

| Measure | Definition | Data Source |
|---|---|---|
| Poverty Rate | % population earning below the poverty income threshold | ACS, 2018 5-year |
| Per Capita Income | Per capita income in the past 12 months | ACS, 2018 5-year |
| % White | % population with race identified as white alone | ACS, 2018 5-year |
| % Black | % population with race identified as Black or African American alone | ACS, 2018 5-year |
| % Hispanic | % population with ethnicity identified as of Hispanic or Latinx origin | ACS, 2018 5-year |
| % No high school diploma | % population 25 years and over, less than a high school diploma | ACS, 2018 5-year |
| % Children | % population under 18 | ACS, 2018 5-year |
| % Vacant housing | % of vacant housing units | ACS, 2018 5-year |
| % Long-term occupancy | % of population who moved into their current housing more than 20 years ago | ACS, 2018 5-year |
| Foreclosure rate | Estimated % of mortgages to start foreclosure process or be seriously delinquent during the 2008 Recession. | Neighborhood Stabilization Program (NSP2), U.S. Department of Housing and Urban Development (HUD), Office of Policy Development and Research, 2009 |
| Traffic volume | Logged total annual average daily traffic counts per road segment. The final result is then scaled from 0 to 100, with higher numbers correspond to greater traffic. | Illinois Department of Transportation (IDOT), 2019 |
| Property crime per 1000 | Property crimes per 1000 residents. Based on the Chicago Police Department, property crimes include arson, burglary, motor vehicle theft and theft. | Chicago Data Portal, 2019. |
| Violent crime per 1000 | Violent crimes per 1000 residents. Based on the Chicago Police Department, violent crimes include assault, battery, criminal sexual assault, homicide, robbery, human trafficking. | Chicago Data Portal, 2019. |

**eTable 2:** Risk activity spaces: within and outside Cook and Collar counties.

| | Buy drugs | | Inject drugs | | Meet sex partners | |
|---|---|---|---|---|---|---|
| | Within boundary | Outside boundary | Within boundary | Outside boundary | Within boundary | Outside boundary |
| suburban | 235 | 1 | 162 | 2 | 83 | 4 |
| transient | 359 | 2 | 233 | 4 | 112 | 20 |
| urban | 422 | 0 | 298 | 0 | 133 | 6 |

***Note.*** Boundary refers to the geographic boundary of Cook & Collar counties.

**eTable 3.** Table 2 in the manuscript with numbers of missing reported.

| Variable | All, N = 258 | Suburban, N = 72 | Transient, N = 77 | Urban, N = 109 | p-value |
|---|---|---|---|---|---|
| Race/Ethnicity | | | | | .025 |
| Hispanic | 65 (25%) | 23 (32%) | 15 (19%) | 27 (25%) | |
| Non-Hispanic White | 153 (59%) | 45 (62%) | 50 (65%) | 58 (53%) | |
| Other | 40 (16%) | 4 (5.6%) | 12 (16%) | 24 (22%) | |
| Gender | | | | | .8 |
| Male | 185 (72%) | 49 (68%) | 56 (73%) | 80 (73%) | |
| Female | 72 (28%) | 23 (32%) | 21 (27%) | 28 (26%) | |
| Transgender | 1 (0.4%) | 0 (0%) | 0 (0%) | 1 (0.9%) | |
| Age | | | | | .3 |
| 17-25 | 48 (19%) | 17 (24%) | 14 (18%) | 17 (16%) | |
| 26-29 | 114 (44%) | 36 (50%) | 33 (43%) | 45 (41%) | |
| 30-34 | 49 (19%) | 10 (14%) | 18 (23%) | 21 (19%) | |
| 35 + | 47 (18%) | 9 (12%) | 12 (16%) | 26 (24%) | |
| Homeless | 167 (65%) | 21 (29%) | 66 (86%) | 80 (73%) | <.001 |
| Homeless (# of days) | 90 (30, 178) | 40 (20, 100) | 78 (30, 90) | 120 (60, 180) | <.001 |
| Unknown | 91 | 51 | 11 | 29 | |

**eTable 4.** Table 3 in the manuscript with numbers of missing reported.

| Variable | All,<br>N = 258 | Suburban,<br>N = 72 | transient,<br>N = 77 | Urban,<br>N = 109 | p-value |
|---|---|---|---|---|---|
| Used crack | 215 (84%) | 58 (82%) | 69 (90%) | 88 (81%) | .2 |
| Unknown | 1 | 1 | 0 | 0 | |
| Injected crack | 101 (39%) | 20 (28%) | 36 (47%) | 45 (41%) | .05 |
| Injected speedball | 112 (43%) | 24 (33%) | 35 (45%) | 53 (49%) | .12 |
| Used methamphetamine | 72 (28%) | 14 (20%) | 25 (32%) | 33 (30%) | .2 |
| Unknown | 1 | 1 | 0 | 0 | |
| Injected methamphetamine | 59 (23%) | 8 (11%) | 25 (32%) | 26 (24%) | .008 |
| Receptive syringe sharing | 106 (42%) | 30 (42%) | 36 (47%) | 40 (38%) | .5 |
| Unknown | 7 | 1 | 1 | 5 | |
| Sharing cookers | 179 (72%) | 50 (70%) | 63 (84%) | 66 (63%) | .011 |
| Unknown | 8 | 1 | 2 | 5 | |
| Backloading | 95 (38%) | 19 (27%) | 40 (53%) | 36 (35%) | .004 |
| Unknown | 7 | 1 | 1 | 5 | |
| Experienced overdose past 6 months | 93 (36%) | 21 (29%) | 36 (47%) | 36 (33%) | .057 |
| Revived with Naloxone past 6 months | 81 (31%) | 17 (24%) | 31 (40%) | 33 (30%) | .086 |
| Months since last overdose | 7 (2, 25) | 12 (3, 28) | 4 (2, 19) | 7 (2, 26) | .06 |
| Unknown | 61 | 12 | 15 | 34 | |
| Months since last Naloxone | 8 (2, 27) | 12 (5, 27) | 5 (2, 25) | 6 (2, 27) | .14 |
| Unknown | 80 | 19 | 18 | 43 | |
| HCV positive | 77 (32%) | 9 (13%) | 22 (29%) | 46 (45%) | <.001 |
| Unknown | 14 | 5 | 2 | 7 | |
| HIV positive | 3 (1.2%) | 0 (0%) | 2 (2.7%) | 1 (1.0%) | .5 |
| Unknown | 13 | 4 | 2 | 7 | |

**eTable 5.** Table 4 in the manuscript with missing value reported.

| Variable | All, N = 258 | Suburban, N = 72 | Transient, N = 77 | Urban, N = 109 | p-value |
|---|---|---|---|---|---|
| | | Injection Network | | | |
| Network degree | 3.00 (2.00, 5.00) | 3.00 (2.00, 4.00) | 4.00 (3.00, 6.00) | 3.00 (2.00, 5.00) | .002 |
| Unknown | 1 | 0 | 0 | 1 | |
| Mean strength of ties | 3.00 (2.75, 3.60) | 3.00 (2.84, 3.62) | 3.00 (2.71, 3.33) | 3.00 (2.66, 4.00) | .5 |
| Unknown | 1 | 0 | 0 | 1 | |
| Age standard deviation | 5.9 (3.5, 8.7) | 3.6 (1.5, 6.5) | 7.1 (4.6, 9.3) | 6.1 (4.3, 9.1) | <.001 |
| Unknown | 36 | 9 | 6 | 21 | |
| Average age of alters | 32 (29, 36) | 31 (27, 35) | 33(30, 36) | 33 (30, 37) | .004 |
| Unknown | 1 | 0 | 0 | 1 | |
| Max age of alters | 38 (32, 47) | 34 (29, 40) | 40 (35, 48) | 40 (32, 50) | < .001 |
| Unknown | 1 | 0 | 0 | 1 | |
| % alters living in Cook | 100 (50, 100) | 50 (0, 100) | 100 (60, 100) | 100 (97, 100) | <.001 |
| Unknown | 1 | 0 | 0 | 1 | |
| Network effective size | 2.06 (1.25, 3.05) | 1.75 (1.32, 2.48) | 2.71 (1.84, 4.00) | 2.00 (1.00, 2.93) | .002 |
| Unknown | 1 | 0 | 0 | 1 | |
| Network tie density | 0.93 (0.73, 1.00) | 1.00 (0.83, 1.00) | 0.86 (0.69, 1.00) | 0.97 (0.80, 1.00) | .014 |
| Unknown | 1 | 0 | 0 | 1 | |
| | | Sexual Network | | | |
| Network degree | 1.00 (1.00, 2.00) | 1.00 (1.00, 1.00) | 1.00 (1.00, 2.00) | 1.00 (1.00, 2.00) | .015 |
| Unknown | 53 | 14 | 10 | 29 | |
| Mean strength of ties | 3.67 (3.00, 4.00) | 4.00 (3.00, 4.00) | 3.33 (2.83, 4.00) | 3.33 (2.50, 4.00) | .070 |
| Unknown | 53 | 14 | 10 | 29 | |
| Age standard deviation | 4.2 (2.1, 7.6) | 2.8 (1.9, 5.2) | 4.7 (3.6, 7.6) | 3.5 (2.1, 7.9) | .4 |
| Unknown | 186 | 60 | 49 | 77 | |
| Average age of alters | 29 (25, 34) | 26 (24, 30) | 29 (25, 33) | 31 (27, 38) | <.001 |
| Unknown | 57 | 15 | 11 | 31 | |
| Max age of alters | 30 (26, 36) | 28 (24, 32) | 30 (26, 36) | 32 (28, 44) | <.001 |

| Variable | All,<br>N = 258 | Suburban,<br>N = 72 | Transient,<br>N = 77 | Urban,<br>N = 109 | p-value |
|---|---|---|---|---|---|
| Unknown | 57 | 15 | 11 | 31 | |
| % alters living in Cook | 100 (0, 100) | 0 (0, 100) | 100 (0, 100) | 100 (63, 100) | <.001 |
| Unknown | 53 | 14 | 10 | 29 | |
| Network effective size | 1.00<br>(1.00, 1.70) | 1.00<br>(1.00, 1.00) | 1.00<br>(1.00, 2.00) | 1.00<br>(1.00, 2.00) | .015 |
| Unknown | 53 | 14 | 10 | 29 | |
| Network tie density | 1.00<br>(1.00, 1.00) | 1.00<br>(1.00, 1.00) | 1.00<br>(0.83, 1.00) | 1.00<br>(0.88, 1.00) | .10 |
| Unknown | 53 | 14 | 10 | 29 | |
| Support Network | | | | | |
| Network degree | 2.00<br>(1.00, 3.00) | 2.00<br>(1.00, 3.00) | 2.00<br>(1.00, 3.00) | 2.00<br>(1.00, 2.00) | .2 |
| Unknown | 12 | 0 | 1 | 11 | |
| Mean strength of ties | 3.50<br>(3.00, 4.00) | 3.67<br>(3.46, 4.00) | 3.33<br>(2.67, 4.00) | 3.58<br>(3.00, 4.00) | .012 |
| Unknown | 12 | 0 | 1 | 11 | |
| Age standard deviation | 13 (4, 19) | 15 (4, 20) | 15 (8, 19) | 8 (3, 15) | .015 |
| Unknown | 116 | 27 | 31 | 58 | |
| Average age of alters | 40 (30, 49) | 40 (31, 48) | 43 (32, 52) | 36 (30, 47) | .11 |
| Unknown | 16 | 1 | 2 | 13 | |
| Max age of alters | 50 (32, 59) | 55 (34, 61) | 54 (36, 59) | 40 (30, 54) | .009 |
| Unknown | 16 | 1 | 2 | 13 | |
| % alters living in Cook | 100 (0, 100) | 18 (0, 100) | 100 (0, 100) | 100 (50, 100) | <.001 |
| Unknown | 12 | 0 | 1 | 11 | |
| Network effective size | 1.00<br>(1.00, 1.75) | 1.12<br>(1.00, 1.67) | 1.00<br>(1.00, 1.68) | 1.00<br>(1.00, 1.99) | .4 |
| Unknown | 12 | 0 | 1 | 11 | |
| Network tie density | 1.00<br>(1.00, 1.00) | 1.00<br>(1.00, 1.00) | 1.00<br>(1.00, 1.00) | 1.00<br>(1.00, 1.00) | .5 |
| Unknown | 12 | 0 | 1 | 11 | |

**eTable 6.** Table 5 in the manuscript with numbers of missing reported.

| Variable | Census tracts in CRASs, N = 51 | Other census tracts in Chicago, N = 747 | p-value |
|---|---|---|---|
| Poverty Rate | 37 (31, 48) | 18 (10, 27) | <.001 |
| Per Capita Income | 13,949 (12,427, 15,978) | 26,421 (18,586, 43,094) | <.001 |
| % White | 5 (2, 11) | 54 (10, 77) | <.001 |
| % Black | 90 (81, 95) | 8 (2, 81) | <.001 |
| % Hispanic | 5 (2, 15) | 13 (4, 43) | <.001 |
| % No high school diploma | 23 (19, 30) | 13 (6, 23) | <.001 |
| % Children (under 18) | 30 (25, 33) | 21 (16, 26) | <.001 |
| % Vacant housing | 19 (15, 24) | 11 (7, 16) | <.001 |
| % Long-term occupancy | 25 (16, 29) | 22 (14, 32) | >.9 |
| Foreclosure rate | 23 (22, 25) | 12 (7, 21) | <.001 |
| Unkown | 19 | 223 | |
| Traffic volume | 5.00 (4.50, 5.38) | 5.00 (4.60, 5.50) | .3 |
| Unkown | 1 | 42 | |
| Property crime per 1000 | 46 (31, 59) | 23 (14, 38) | <.001 |
| Violent crime per 1000 | 88 (74, 110) | 18 (11, 47) | <.001 |

***Note.*** Median (IQR) was provided for each variable. P-value was calculated with Wilcoxon rank sum test.

**eTable 7.** Average travel distance (miles) for different activities per residential group.

| Activity | Overall, N = 258 | Suburban, N = 72 | Transient, N = 77 | Urban, N = 109 | p-value |
|---|---|---|---|---|---|
| Drug purchasing | 6.03 (3.41, 14.50) | 13.51 (5.94, 21.34) | 10.84 (6.92, 37.22) | 3.62 (1.38, 5.12) | <0.001 |
| Unknown | 4 | 2 | 1 | 1 | |
| Drug injection | 4.91 (1.85, 11.60) | 7.25 (2.85, 16.64) | 9.89 (5.92, 37.00) | 2.44 (1.01, 4.05) | <0.001 |
| Unknown | 7 | 3 | 2 | 2 | |
| Meet sex partners | 6.75 (2.67, 16.55) | 6.92 (2.99, 17.51) | 13.43 (8.02, 51.24) | 3.16 (0.96, 6.74) | <0.001 |
| Unkown | 45 | 12 | 11 | 22 | |

***Note.*** Median (IQR) was provided for each variable. P-value was calculated with Kruskal-Wallis rank sum test.

More information regarding KDE:

There are two important considerations when implementing KDE analyses, including the actual kernel used to weight the point locations of each activity and the bandwidth ($h$) of observations to be weighted by the kernel. One could specify different kernels as long as the selected one is symmetric and represents a continuous probability density function with a mean of zero and a bounded variance, for which popular choices are the Gaussian and Epanechnikov. We tried several kernels and found no substantial differences in the results. We thus presented our results with the widely used Gaussian smoothing. The second consideration regarding the bandwidth is more relevant as it determines the direction and the amount of smoothing. We explored several alternatives for the bandwidth ($h$) using Least Square Cross Validation (LSCV) schemes and ad-hoc criteria. Some of the LSCVs did not converge, but the ad-hoc criteria provided similar estimates to the ones obtained by the best LSCV approximations. As such, we reported the results using the ad-hoc criteria. The bandwidth $h$ was obtained based on Equation (1), where $\sigma$ represents the average variance of points across longitude ($x$) and latitude ($y$), and $n$ refers to the number of point locations for each activity-group pair (e.g., drug injection for urban group). Having obtained the KDEs, ranges of activities were derived using KDE contours (the function gettverticeshr was employed). Based on the ranges of activities, we identified concentrated risk activity spaces (CRAS) within Cook and Collar counties where most activities took place for each residential group.

$$h = \sigma \cdot n^{-\frac{1}{6}}; \ \sigma^2 = 0.5 \cdot (var(x) + var(y)) \quad (1)$$
